# Polygenic Risk and Genetic Predisposition in Post-Traumatic Epilepsy: A Framework for Risk Estimation

**DOI:** 10.64898/2026.09.10.26362755

**Authors:** James W.Y. Chen, Cindy Le, Kathleen Cui, Olga Alexeeva, Janice Joo, Julia Bailey

## Abstract

**Background:** Post-traumatic epilepsy (PTE) can be a lifelong complication of traumatic brain injury (TBI). We hypothesize that PTE develops according to a Two-Hit Hypothesis, in which the first “hit” is a genetic predisposition and the second “hit” is the TBI. Using two independent datasets, we provide the first evidence that PTE is a polygenic disorder, and we propose a whole-exome sequencing (WES) based framework to estimate the risk of developing PTE.

**Methods:** From a pool of thousands of screened veterans, we recruited a cohort of PTE subjects (n=28) and a control cohort of TBI subjects without PTE (n=22) and then performed WES. Approximately 375,000 variants identified in each subject were compared to approximately 15,000 verified epilepsy-associated variants from ClinVar. Fisher’s exact test was used to identify variants associated with PTE, which were subsequently classified as either PTE-prone or PTE-protective. Odds ratios (ORs) were calculated at both the variant and subject levels. Receiver operating characteristic (ROC) curve analysis was used to evaluate the predictive model. A second dataset was used to validate the model, and logistic calibration was performed to estimate the probability of developing PTE.

**Results:** Thirty PTE-prone and 56 PTE-protective variants were identified, with corresponding large-effect ORs. ROC analysis demonstrated excellent discrimination (AUC=0.97). Polygenic variant patterns differed between cohorts, with a predominance of PTE-prone variants in affected individuals and PTE-protective variants in controls.

**Conclusion:** Our findings support a polygenic framework consistent with the Two-Hit Hypothesis. In addition, we developed a framework to estimate individual polygenic risk for PTE.

## Introduction

Epilepsy is a disorder of the brain characterized by recurrent unprovoked seizures and is frequently accompanied by neurobiological, cognitive, psychological, and social comorbidities^1^. Post-traumatic epilepsy (PTE), traditionally categorized as an acquired epilepsy without a known genetic predisposition, is a well-recognized and often lifelong complication of traumatic brain injury (TBI)^2–4^.

Between 1961 and 2015, approximately 20 international epidemiological studies of varying scales and methodologies have investigated risk factors for PTE after TBI^5^. These studies identified several factors associated with increased risk, including male gender, loss of consciousness at the time of TBI, post-traumatic amnesia, focal neurologic deficits, early post-traumatic seizures (occurring more than one week after TBI), age over 65 years, and prior history of alcohol abuse. The relative odds ratios (ORs) for these risk factors are estimated to range from 1.31 to 2.18. Greater injury severity and abnormal neuroimaging findings—such as skull fracture, acute intracerebral hematoma, subdural hemorrhage, or cerebral contusion—are associated with an even greater risk of PTE, with reported odds ratios ranging from 2.27 to 2.65.

Among military personnel with combat-related TBI, the incidence of PTE has historically ranged from 35 to 45 percent across conflicts including World War I, World War II, and the Korean War^2,6^.

Findings from the Vietnam Head Injury Study further underscore this high burden: approximately 53 percent of veterans with penetrating head injuries experienced at least one seizure□. During the Korean War, no specific prophylactic treatments were provided to veterans. Meanwhile, during the Vietnam War, military personnel with a penetrating head injury received phenytoin for six months, with no apparent reduction in the incidence of PTE^2^. Consistent with these findings, multiple large clinical trials have failed to demonstrate the ability of antiseizure medications to prevent epileptogenesis in PTE^2^.

In approximately half of patients, the first seizure occurs within one year of TBI; however, nearly 15 percent of patients develop PTE more than 5 years after the initial injury^8,9^. A large cross-sectional analysis of the Veterans Affairs fiscal years 2009–2010 database (including 256,284 veterans) estimated the prevalence of epilepsy to be 10.6 per 1,000 veterans of Operations in Afghanistan and Iraq. Epilepsy was strongly associated with a prior diagnosis of penetrating TBI (adjusted OR: 18.77; 95% confidence interval: 9.21–38.23). Other forms of TBI were also associated with increased risk of PTE (OR: 1.64; 95% CI: 1.43–1.89), with even mild TBI thought to substantially increase the likelihood of developing PTE^1^□. The study also identified statistically significant associations between epilepsy and several other conditions, including schizophrenia, bipolar disorder, headaches, and substance use disorders.

### The Two-Hit Hypothesis

Similar to the two-hit hypothesis in oncology^11–13^, we propose that the development of PTE requires a genetic predisposition constituting the first hit, followed by subsequent TBI incurring the second hit, thereby initiating epileptogenesis. We hypothesize that mild/moderate TBIs are more likely to activate larger-effect genes, whereas severe TBIs are more likely to activate many more smaller-effect genes. Our results support this two-hit hypothesis.

We also develop a pioneering framework to estimate the genetically predisposed risk of PTE from the first hit. This variants–based calculator can be applied either before or after TBI.

## Methods

Using a case-control design, we screened 2,378 veterans from the VAGLAHS and associated VA sites in Southern California. We then recruited a cohort of PTE patients and another control cohort of TBI patients without epilepsy. Participants were grouped in two batches: the first was used to develop the model (TBI-without-PTE, n=22 vs. PTE, n=28), and the second was used to test it (TBI-without-PTE, n=24 vs. PTE, n=3). PTE subjects had to 1) have a diagnosis of PTE confirmed by an epileptologist, and 2) have had seizure onset >1 month and <10 years after TBI (**Table 1**, with a median of 1 year and a mean of 3 years). Individuals with a different probable etiology of epilepsy, such as genetic epilepsy or a positive family history of epilepsy, were excluded to avoid including genetic epilepsy of Mendelian inheritance. The clinical criteria of mild, moderate, and severe TBI were applied^2^. To ensure comparability with respect to injury severity, most participants in both cohorts had mild TBIs (Table 1: 75% in the PTE cohort and 73% in the TBI cohort). The cohorts were compatible with respect to demographic characteristics, including age at injury and at recruitment (Table 1). Exclusion criteria for both cohorts included having an active mood or psychotic disorder within the past year, history of illicit substance use, and history of alcohol abuse. Subjects with any major neurological disorder or abnormal brain MRI findings were excluded. The TBI control cohort had a median of 17 years and a mean of 18 years from the last TBI to recruitment, suggesting that the probability of including false-negative PTE cases was low (**Table 1**). All subjects consented to the study by signing a VAGLAHS-approved IRB protocol form and a HIPAA form for PHI protection. The IRB protocol was approved by the CDMRP prior to subject recruitment.

**Table 1.** Demographics of PTE and TBI Cohorts for the model. There are 28 subjects in the PTE cohort and 22 in the TBI cohort. The severity of TBIs and demographics are compatible between the two cohorts. In the PTE cohort, the mean duration from the last TBI to seizure onset was 2.5 years, and the median time was 1 year. In the TBI cohort, the mean time from the last TBI to sample collection was 17.6 years, which is 7-fold that of the average time to seizure onset in the PTE cohort. This shows that sufficient time has passed to separate the two cohorts, and that the likelihood of a false-negative PTE subject being recruited into the TBI cohort is low.

|  | **<u>PTE Cohort</u>** | **<u>TBI Cohort</u>** |
| --- | --- | --- |
| **Gender** | 2 (7%) female, 26 (93%) male | 3 (13.6%) female, 19 (86.3%) male |
| **Ethnicity** | 22 Caucasian (78.5%)<br>5 African American (17.8%)<br>1 American Indian (3.5%) | 18 Caucasian (81.8%)<br>2 African American (9.1%)<br>2 American Indian (9.1%) |
| **Severity of TBI** | 75% mild, 25% moderate | 73% mild, 18% moderate, 9% severe |
|  | **<u>Median, Mean, min, max yrs</u>** | **<u>Median, Mean, min, max yrs</u>** |
| **Age at time of recruitment** | 46, 49 (25 – 75) | 42, 45 (32 – 75) |
| **Age of TBI** | 25, 26 (13 – 66) | 24, 25 (14 – 46) |
| **Time from Last TBI to seizure onset** | 1, 3 (1 – 10) | N/A |
| **Last TBI to time of collection** | 20, 22 (1 – 46) | 17, 18 (2 – 54) |
| **First TBI to time of collection** | 20, 22 (1 – 46) | 18, 20 (2 – 54) |
| **Total subjects** | 28 | 22 |

DNA was extracted from either a fresh blood sample or a saliva kit (OGR-500, DNA Genotek Inc.). Samples were preserved in a −80°C freezer and submitted in two separate batches to the Center for Inherited Disease Research at Johns Hopkins University (Genetic Resources Core Facility; RRID: SCR_018669) for whole-exome sequencing (WES). The first batch was sequenced at the end of 2024 to build the PTE risk estimation model. The second batch was sequenced at the beginning of 2026 to test the model. On October 2, 2024, a template of approximately 15,000 epilepsy-related variants was constructed from the ClinVar database using criteria of multiple submitters and expert review. Approximately 375,000 variants were identified from each subject’s WES and then matched against this template to create a dataset of matches per subject. Fisher’s exact test was used to compare the PTE and TBI cohorts and identify variants associated with PTE (defined by p-value ≤0.05), with separate analyses for recessive or dominant inheritance assumptions for each variant.

For each PTE-related variant, contingency tables were used to calculate the OR for developing PTE. Variants were classified as PTE-prone (OR>1) or PTE-protective (OR<1). After converting to the natural logarithm scale, the total ln (OR)s per subject was obtained by combining the PTE-prone and PTE-protective ln(OR)s. ROC curves with AUC were calculated to evaluate the model’s discriminative ability for predicting PTE.

To convert total ln(OR)s to the probability of developing PTE, logistic calibration was performed in MATLAB using the function glmfit to obtain the intercept (α) and slope (β), and the fitted logistic regression curve using equation (1). The probability (P) of developing PTE is calculated using the inverse equation (2).

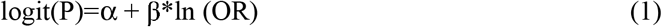

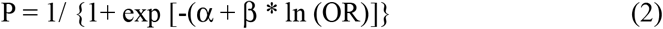

To test the risk estimation model, the second batch of WES data was analyzed using the same protocol to evaluate the framework’s predictive performance. All the data processing and statistical analysis were conducted using MATLAB and MS Excel.

## Results

From the first dataset, we identified 30 PTE-prone and 56 PTE-protective genetic variants (p≤0.05). The OR for each identified variant was computed (**Table 2**). Visual comparison of the two cohorts showed that the PTE cohort contained a greater proportion of PTE-prone variants (**Figure 1**, pink region), whereas the TBI cohort contained a greater proportion of PTE-protective variants (**Figure 1**, green region). This difference was statistically significant when comparing the means (**Figure 1**, legends). The total number of variants was significantly different between the two cohorts, with more total variants in the TBI-without-epilepsy cohort due to an overwhelming predominance of PTE-protective variants. In the second dataset, there was a similar pattern as the first but with one exception: no statistical difference in the PTE-prone variants between the two cohorts was identified, likely due to sampling error from the low number of PTE cases (n = 3). Our data suggest that the number of PTE-protective variants is probably the predominant factor influencing phenotypic manifestation.

**Table 2.**
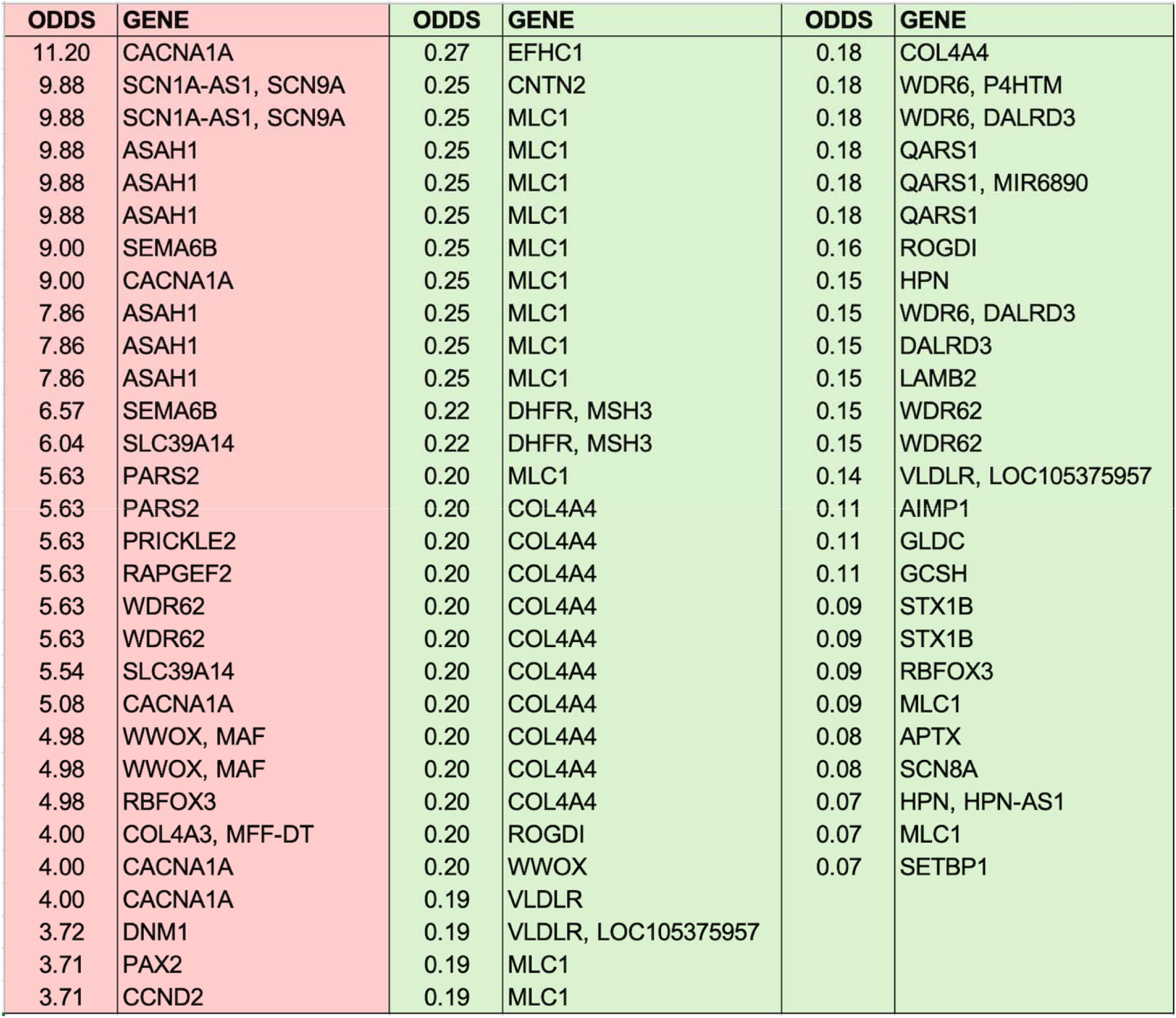
The ORs of PTE-prone variants (listed by genes), highlighted in pink, and PTE-protective variants (listed by genes), highlighted in green. Repeated items are due to multiple variants in the gene.

**Figure 1.**
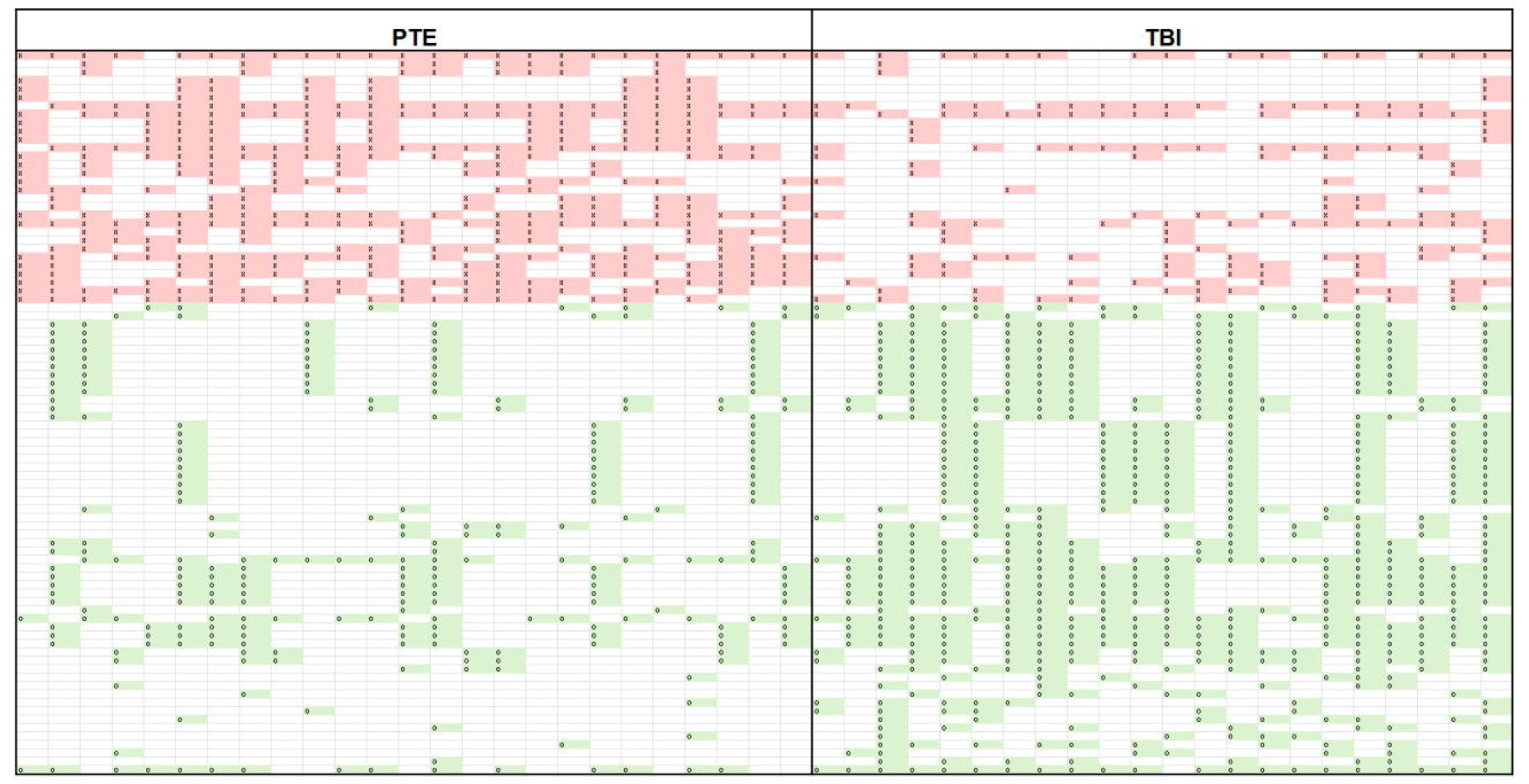
This figure shows the distribution of polygenic variants across all subjects in both cohorts from the first dataset (2024 sequenced). Each column represents a subject. Each row represents a specific variant across the subjects. The pink-highlighted cell represents a PTE-prone variant (marked by “x”), and a green-highlighted cell represents a PTE-protective variant (marked by “o”). The differences between the two cohorts are apparent on visual inspection, with a higher proportion of PTE-prone variants in the PTE cohort and a higher proportion of PTE-protective variants in the TBI cohort. Statistical analysis was performed and listed below with mean ± 1.96 x sem, which is equivalent to 95% confidence interval, for the counts of respective types of variants: 2024 PTE cohort, total variants: 27.4 ± 2.8, 2024 PTE cohort, PTE-prone variants: 16.6 ± 2.9, 2024 PTE cohort, PTE-protective variants: 10.8 ± 2.9; 2024 TBI cohort, total variants 36.5 ± 4.9, 2024 TBI cohort, PTE-prone variants: 7.6 ± 1.3; 2024, 2024 TBI cohort, PTE-protective variants: 28.9 ± 4.5; For comparison (the variant map not shown), the statistical analysis of the second dataset shows: 2026 PTE cohort, total variants: 25.3 ± 5.1, 2026 PTE cohort, PTE-prone variants: 14 ± 1.3, 2026 PTE cohort, PTE-protective: 11.3 ± 4.0; 2026 TBI cohort, total variants: 33.7 ± 3.3, 2026 TBI cohort, PTE-prone variants: 12.4 ± 1.4, 2026 TBI cohort, PTE-protective variants: 21.3 ± 2.9.

The histograms of total ORs in the respective cohorts were plotted in different colors: blue for the TBI cohort and orange for the PTE cohort (**Figure 2A**). The distributions were distinct but partially overlapping. ROC curve analysis (**Figure 2B**) yielded an AUC of 97%, indicating strong discriminative ability. When the model was evaluated with the second dataset, the results were consistent with those of the initial analysis: histogram distributions were comparable (**Figure 2C**), and ROC analysis combining both datasets (**Figure 2D**) yielded an AUC of 94%.

**Figure 2.**
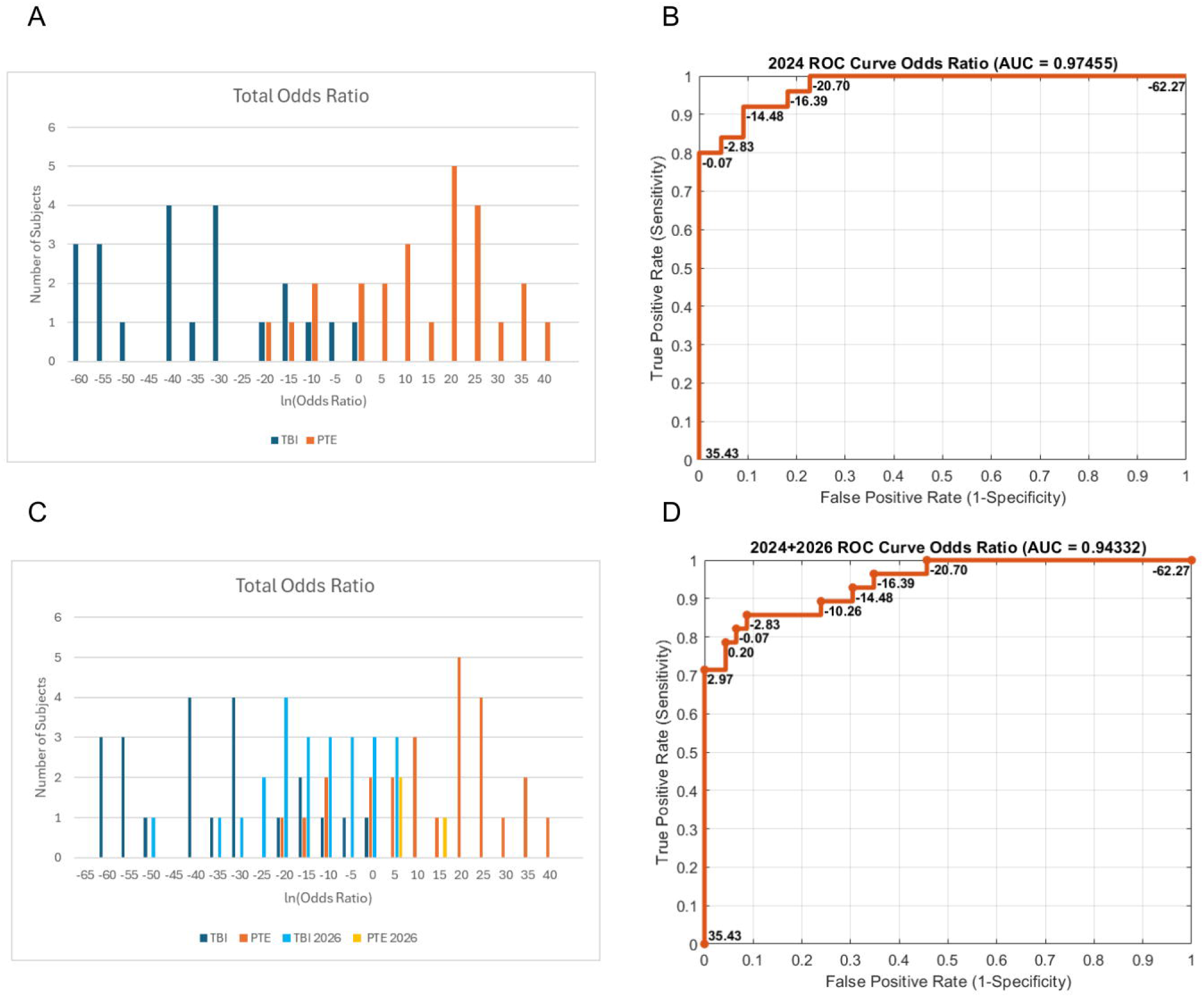
2 (A): It shows the histogram of the logarithm of the ORs in the TBI cohort in blue and the PTE cohort in orange. 2 (B): It represents the ROC curve analysis using the dataset from 2 (A), which results in an AUC of 97%. 2 (C): It shows the histogram of 2 (A), which is superimposed with the second dataset (data sequenced in 2026), with the new TBI cohort in light blue and the PTE cohort in yellow. 2(D) The ROC curve analysis was performed using combined data from 2 (C), resulting in an AUC of 94%.

To convert the logarithm of the total ORs to the probability of developing PTE, logistic calibration was performed on the model, yielding an intercept of 2.19 and a slope of 0.16 of the fitted regression curve (**Figure 3**). The arbitrary thresholding values can be determined using two different methods: 1) the average of the means of the PTE and TBI-without-epilepsy cohorts, which is a probability of 0.5 (green dot); or 2) maximizing the (true positive rate – false positive rate), which is at a probability of 0.45 (blue dot). According to the model, the risk of developing PTE is negligible when the calculated probability is below 0.2, and almost certain when the calculated probability is 0.9 or higher.

**Figure 3.**
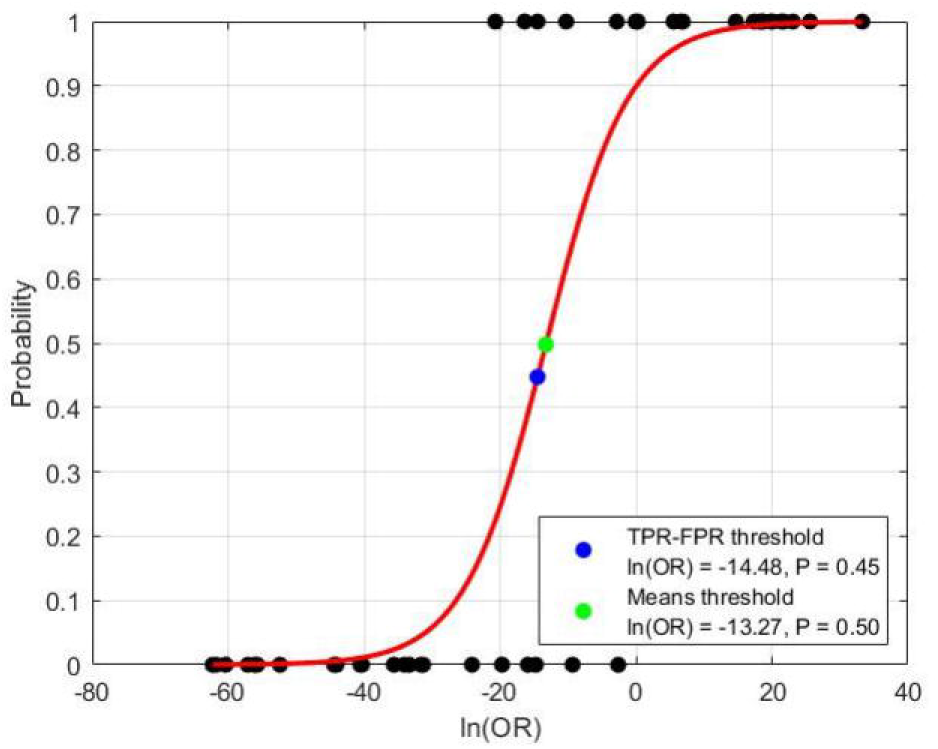
This figure is the logistic calibration of the logarithm of the ORs to obtain the probability of developing PTE. Two commonly used thresholding values are marked with a green dot (the average of the means of the two cohorts) at probability = 0.5, and a blue dot (maximizing (TPR – FPR)) at probability = 0.45.

## Discussions

PTE has traditionally been categorized as an acquired epilepsy caused by TBI^1,2^. Our findings challenge the traditional view that PTE is acquired, instead supporting a genetic predisposition hypothesis with a polygenic form. These results are consistent with the two-hit hypothesis, as well as with recent shifts in opinion^14–16^. Using this new paradigm, several observations about PTE can be better understood. For instance, it was previously noted that only about 50% of subjects developed PTE after a penetrating brain injury^2^. This proportion could be consistent with our findings, which suggest that probably only about 50% of the general population is genomically predisposed to develop PTE after severe TBI. Similarly, clinical trials of prophylactic antiseizure medications after TBI have failed to prevent PTE in large, controlled studies^2^. Per this paradigm, the inability of antiseizure medications to prevent PTE in these studies may have been explained by the comparable distributions of PTE polygenic risk across large experimental and control cohorts. The polygenic framework may also explain why the high frequency of mild TBI among veterans increases epilepsy prevalence only modestly, which showed a ceiling effect to a degree still within community ranges^10^. Our findings support the crucial role played by genomic predisposition in PTE development following the two-hit hypothesis^11–13^.

We constructed a variant filter using the ClinVar database that, while unorthodox, produced an efficient filter for removing unwanted variants. In electrophysiology, due to the need for real-time noise reduction (e.g., removing muscle artifact from EEG) in time-series data, the default approach is to apply a specific filter^17^ rather than to perform statistical analysis such as logistic regression with multiple-testing corrections. Although filtering can introduce minor signal distortion, this approach improves the signal-to-noise ratio significantly and is straightforward to implement. While WES variants are not time-series data, they can be analyzed similarly by applying an acceptable filter to reduce data noise. This approach enabled identification of ClinVar-verified epilepsy variants within each subject, allowing for at least 96% of SNP variants to be filtered out. With p≤0.05 set in Fisher’s exact test, only 86 variants were identified out of the 15K ClinVar variants, representing approximately 0.6% of the ClinVar template and 0.02% of the 375K variants identified per subject by WES.

In theory, testing 15,000 variants at p≤0.05 could yield about 750 false positives; it was nearly an order of magnitude greater than the 86 variants detected in our cohorts. Moreover, the observed effect sizes were large: PTE-prone variants had ORs of 3.71–11.20, whereas PTE-protective variants had ORs of 0.07–0.27 (corresponding to a 3.7–14.3-fold reduction in risk). Thus, although the filtering approach may have introduced minor distortion analogous to electrophysiologic filtering, these large effect variants argue against spurious detection. The second dataset, while smaller than planned due to time constraints during the grant period, showed a consistent pattern of distinct PTE-prone and PTE-protective variant groups, with a predominance of protective variants in the cohort that did not develop PTE after TBI.

One primary advantage of our methodology is its ability to classify variants as PTE-prone or PTE-protective. This binary distinction is crucial to the functional interpretation of these variants’ roles in epileptogenesis and phenotype manifestation. For instance, a PTE-protective variant could be a loss-of-function mutation in cerebral excitatory pathways or a gain-of-function mutation in inhibitory pathways.

Another benefit of this methodology is the potential ability to better understand genotype-phenotype discordance^18^, which has posed a central challenge in clinical genomics and to the genetic epilepsy research community as a whole^19^. Possible explanations have included incomplete penetrance, variable expressivity, polygenic background, environmental influences, and others. Here, we propose a framework to estimate the polygenic risk of PTE, thus providing an operational method to evaluate whether polygenic variation contributes to genotype-phenotype discordance in PTE.

Moving forward, this methodology could enable polygenic disorders to be investigated with much smaller cohorts than those required for genome-wide association studies, significantly reducing costs and accelerating progress in clinical polygenic disorder research. Additionally, identifying large-effect PTE-related variants could enable the development of polygenic animal models to study interactions between genetic susceptibility and environmental factors. In such models, the environmental insult of TBI could be precisely controlled with respect to injury timing, type, and severity, while combinations of PTE-prone and PTE-protective variants could be manipulated experimentally. Although modeling dozens of variants would require very large breeding populations^21,22^, this strategy may ultimately provide a powerful platform for dissecting how environmental factors interact with polygenic predispositions, opening a new road for the understanding of epileptogenesis.

Our risk estimation framework has several areas for future improvement. For situations when multiple variants are identified within a gene, a linkage disequilibrium-based modifier could be incorporated into future iterations of the model once additional molecular data are available to address the potential issue of high linkage disequilibrium^22^. Additionally, as our framework was derived from a population of veterans with un-excluded comorbidities of anxiety disorder, PTSD, and migraine^10^, in its current state, it may be most accurate for predicting the probability of PTE in people with similar traits.

## Data Availability

All data produced in the present study are available upon reasonable request to the authors

## Acknowledgment

Virginia Janovsky, RN, provides study coordination and research protocol maintenance. Technical support was provided by Miyabi Tanaka, M.D., for DNA extractions, Psbhavanishankara Gowda, Ph.D., and Julia Reitkopp for subject screening.

## Funding

CDMRP EP 190044, PI: James W.Y. Chen, M.D., Ph.D.

We confirm that we have read the Journal’s position on issues involved in ethical publication and affirm that this report is consistent with those guidelines.

